# Quantify the Contribution of Modifiable Risk Factors for Progression of MGUS to Multiple Myeloma

**DOI:** 10.1101/2025.04.21.25326164

**Authors:** Mei Wang, Byron Sigel, Lawrence Liu, John H. Huber, Mengmeng Ji, Martin W. Schoen, Kristen M. Sanfilippo, Theodore S. Thomas, Graham A. Colditz, Shi-Yi Wang, Su-Hsin Chang

**Author notes:** Corresponding Author: Su-Hsin Chang, PhD SM Address: 660 S. Euclid Ave. Campus Box 8100, St. Louis, MO 63110. Co-first authors (Mei Wang, Byron Sigel).

## Abstract

**Background:** Multiple myeloma (MM), the most common plasma cell dyscrasia in the U.S., is preceded by an asymptomatic precursor monoclonal gammopathy of undetermined significance (MGUS). Although several risk factors for MGUS progression are known, their relative contributions remain unclear. Unlike other malignancies, such evidence is lacking for MM despite its high burden.

**Objective:** To quantify contributions of modifiable risk factors to MGUS progression to MM to inform prevention.

**Design:** Retrospective cohort study.

**Setting:** Nationwide U.S. Veterans Health Administration (VHA).

**Participants:** Patients with MGUS (IgG, IgA, or light chain) diagnosed from 10/1/1999-12/31/2023.

**Interventions:** Modifiable risk factors including excess body mass index (BMI), chemical exposure, and comorbidities.

**Measurements:** Excess body mass index was defined as BMI ≥25 kg/m², chemical exposure was measured by prior exposure to Agent Orange, comorbidities were summarized using Charlson Comorbidity Index. Multivariable-adjusted population attributable fractions (aPAF) was calculated for each modifiable risk factor. The aPAF estimates the proportion of progression in patients diagnosed with MGUS that could have been prevented, if a given risk factor were absent.

**Results:** The cohort included 35,073 MGUS patients (33,670 [96.0%] male and 23,218 [66.2%] White), of whom 2,895 (8.3%) progressed to MM. Median age at MGUS diagnosis was 71.8 (IQR: 64.4-78.6) years. Among all evaluated risk factors, excess BMI was the leading factor (Black: aPAF=27.1%, 95% CI 19.5-34.0%; White: 27.2%, 95% CI 20.3-33.4%; All: aPAF=27.1%, 95% CI: 22.1-31.9%).

**Limitations:** Potential residual confounding, limited generalizability beyond the VHA population.

**Conclusion:** Our study highlights the potential for weight management as a key strategy in reducing the risk of progression to MM in Black and White patients diagnosed with MGUS.

**Primary Funding Source:** National Institutes of Health.

## INTRODUCTION

Multiple myeloma (MM) is the most prevalent plasma cell malignancy, comprising ∼10% of hematologic cancers.^1,2^ In 2024, there were 35,780 new diagnoses and 12,540 deaths in the U.S.^3^ MM develops from monoclonal gammopathy of undetermined significance (MGUS), a pre-malignant condition present in 3-10% of individuals ≥50 years and twice as common in those of African descent.^4–7^ MGUS carries a 1% annual progression risk to MM or other lymphoproliferative disorders.^5,8^ MM remains incurable, with a 5-year survival rate of 61.1% (2014-2020)^3^ and imposes a significant financial burden.^9,10^ Identifying and quantifying modifiable risk factors for MGUS progression is crucial for risk stratification and targeted interventions.^11,12^

Prior studies have identified potential modifiable risk factors for MM, including chemical exposure,^13–17^ cumulative exposure to overweight/obesity,^18^ and comorbidities.^19–26^ However, their contribution to MM burden remains poorly understood. Addressing this gap is essential for efficient healthcare resource allocation and improved MGUS follow-up.

MM prevention is further complicated by established MM health disparities, where Black individuals bear a higher burden of MM, compared to their White counterparts.^3,26–31^ Understanding the contribution of modifiable risk factors to MM disparities could inform targeted interventions to reduce racial disparities.

We conducted a population-based study to estimate multivariable-adjusted population attributable fractions (aPAFs) overall and by race in the Veteran population. Evidence on risk factors with the highest population attributable risks were identified in other malignancies;^32–38^ however, as one of the highest burdensome cancers, MM is lacking such evidence. By quantifying the contribution of each modifiable risk factor, we identify key intervention targets to reduce MM burden.

## METHODS

### Data and Study Population

Data from the nationwide U.S. Veterans Health Administration (VHA) were used. We selected the Veteran population due to their distinct demographic characteristics, including older age, predominantly male composition, and higher prevalence of Agent Orange (AO) exposure. These factors are associated with an increased risk of MM and progression of MGUS to MM, placing this group at a heightened risk compared to the general population. Chemical exposure, a crucial modifiable risk factor typically assessed through occupational history, is often poorly documented in most databases, including VHA. However, the VHA database provides comprehensive records of AO exposure. AO, an herbicide used during the Vietnam War Era, is contaminated with 2,3,7,8-tetrachlorodibenzo-p-dioxin (TCDD), a highly toxic compound. TCDD remains a prevalent occupational hazard due to its prolonged half-life of ∼3.2 years.^39–43^ Given the well-documented AO exposure in the VHA database, it serves as a reliable proxy for chemical exposure.

The study was approved by Institutional Review Boards at both Veterans Affairs Saint Louis Healthcare System and Washington University School of Medicine.

### Analytic Cohort

We confirmed 50,096 Black/White patients diagnosed with MGUS from 10/1/1999-12/31/2023 with immunoglobulin (Ig) subtype data (eFigure 1). MGUS and MM diagnosis and the corresponding diagnosis date were determined by validated natural language processing (NLP) models.^44^ These models utilized clinical, laboratory, medication, and pathologic data to extract evidence for MGUS and MM achieving 93% accuracy for MGUS and 99% for MM,^44,45^ as compared to only 20% and 58% using billing codes alone.^44,46^

**Figure 1.**
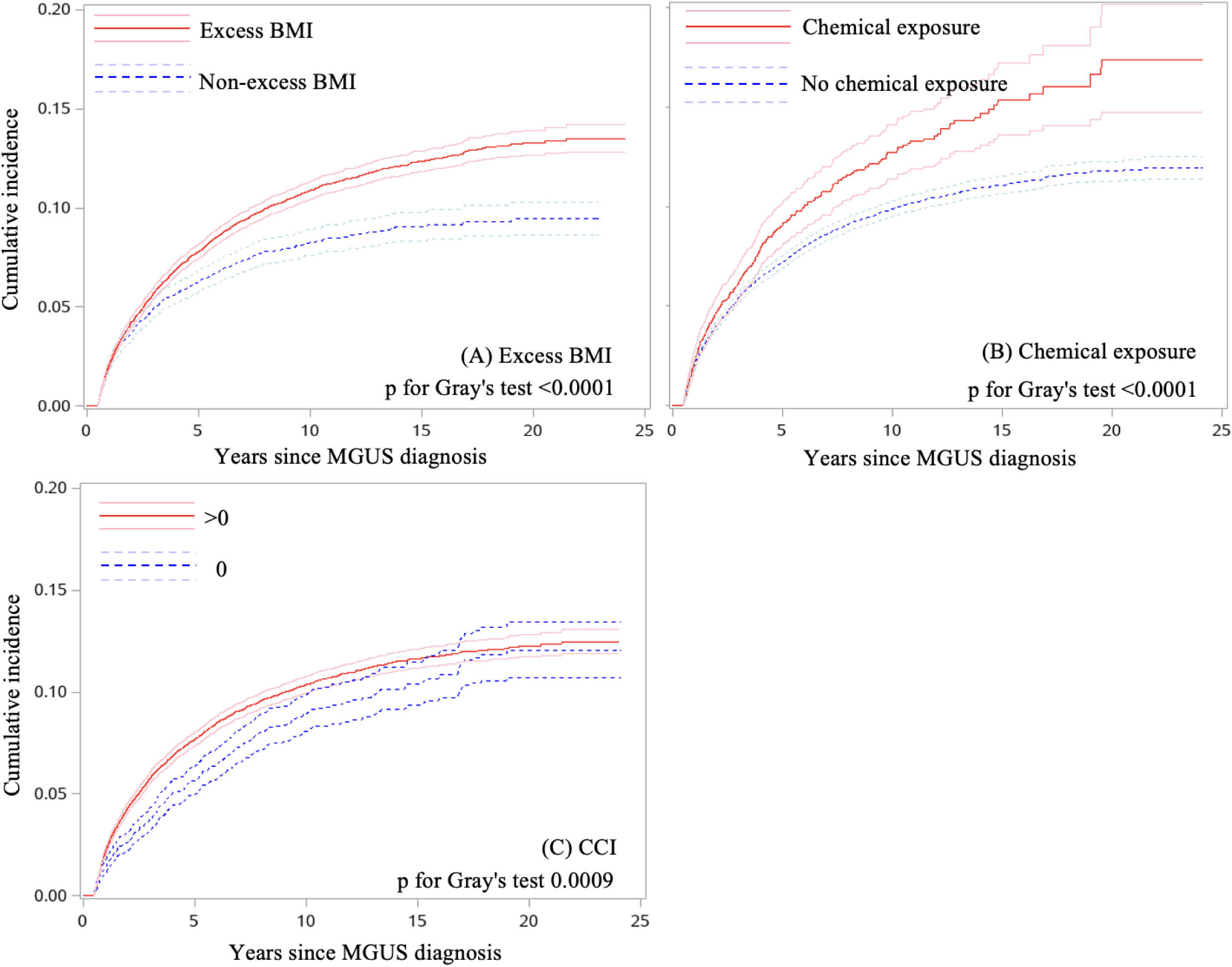
Cumulative incidence of progression to MM among 35,073 MGUS patients for each modifiable risk factor.

Self-reported race was ascertained and recoded as Black/White following published guidelines.^47^ Patients with non-IgG/IgA/light chain subtypes were excluded due to their infrequency and minimal progression risk (n=9,084). Patients progressing <6 months were excluded because they may have had MM at MGUS diagnosis (n=5,898). Finally, patients were excluded if documented date of death was on or before their MGUS diagnosis (n=41). Final analytic cohort consisted of 35,073 patients, among whom 33.8% were Black.

### Outcome Measure

The outcome variable was the progression of MGUS to MM, if any, and the time to progression. MM diagnosis and date were confirmed by the aforementioned NLP model.^44^

### Risk Factors and Select Modifiable Risk Factors to highlight

Multiple data domains were linked to obtain patient-level data on risk factors for progression of MGUS to MM selected based on published work,^6,11,26,48^ clinical judgement, and data availability. These risk factors included older age, male sex, Black race, elevated body mass index (BMI), monoclonal (M-)spike level >1.5 g/dL, IgA subtype, comorbidities (summarized by Charlson Comorbidity Index, CCI^49–51^), and chemical exposure (measured by AO exposure)^42^.

Among these risk factors, BMI, chemical exposure, and CCI are potentially modifiable. BMI was calculated based on the most frequently reported height and weight ≤1 year prior and closest to MGUS diagnosis. Excess BMI was defined as BMI ≥25 kg/m^2^.^48^ Chemical exposure was determined by whether the patient met all of the following criteria: documented AO exposure, AO location designated as Vietnam, and ever served in the Army, Air Force, Navy, or Marine Corps during the Vietnam War Era.^42^ CCI was calculated based on comorbidities present ≤1 year before MGUS diagnosis.^50,51^

### Statistical Analysis

### Patient characteristics

Patient characteristics, stratified by progression status were summarized as proportions for categorical variables and medians (interquartile ranges, IQRs) for continuous variables. Statistical significance for differences across groups was assessed using chi-square tests for categorical variables and Kruskal-Wallis tests for continuous variables.

### Competing risk analyses

Death without progression to MM was considered as a competing event. For each of the studied modifiable risk factors, we plotted the cumulative incidence function (CIF) curves, stratified by individual categories of that risk factor. Gray’s test was performed to compare the cumulative incidence of progression between categories.

Multivariable-adjusted hazard ratios (aHR) and 95% confidence intervals (CI) for each risk factor were estimated using Fine-Gray distribution hazard models.^52^ Follow-up time was defined as time from MGUS to progression, death, or date of censoring (12/31/2023), whichever occurred first. The covariates in the multivariable models included: age, gender (male, female), race (White, Black), Ig subtype (IgG, IgA, light-chain), M-spike level (0, not quantifiable, ≤1.5, >1.5 g/dL, missing), BMI (non-excess <25, excess ≥25 kg/m^2^, missing), all at MGUS diagnosis, and chemical exposure (yes, no), year of MGUS diagnosis, CCI (0, >0). Ig subtype was extracted using a published NLP pipeline.^44^

### aPAF analyses

For each target modifiable risk factor significantly associated with MGUS progression in the multivariable analysis, aPAF was calculated. The aPAF quantifies the proportion of progression events from MGUS to MM, that may have been prevented if the specific risk factor were absent in MGUS patients. It incorporates both the prevalence of each risk factor in the cohort and the adjusted relative risk for MM incidence associated with this risk factor during follow-up. It also accounts for competing risk of death to avoid overestimating aPAFs due to ignoring censoring due to death.^53,54^ We estimated aPAFs for the following risk factor modifications: excess to non-excess BMI, chemical exposure to no chemical exposure, and CCI>0 to CCI=0.

### Race-specific analyses

To assess racial differences in the contribution of modifiable risk factors to the progression of MGUS to MM, the analytic cohort was stratified by race. Differences in aPAFs between the two racial groups were analyzed using two-sample z-tests.

### Sensitivity analyses

We repeated the analyses but replacing excess BMI with BMI classification: underweight <18, normal weight 18-<25, overweight 25-<30, obese ≥30, and missing. We calculated aPAFs for the following modifications: overweight to normal weight, obese to normal weight, chemical exposure to no chemical exposure, and no comorbidities to at least one comorbidity.

All tests were two-sided. Statistical significance was assessed at the level of 0.05. Data queries were performed using SQL Server Management Studio 18 (Microsoft, Redmond, WA). Analyses were performed using SAS 9.3 (SAS Institute Inc., Cary, NC).

## RESULTS

The analytic cohort included 35,073 MGUS patients, of whom 2,895 (8.3%) progressed to MM, and 15,541 (44.3%) died without progression. Median follow-up was 3.8 (IQR: 1.5-7.3) years. The median age of MGUS diagnosis was 71.8 (IQR: 64.4-78.6) years (Table 1). The cohort was predominantly male (96.0%) and White (66.2%), with 73.1% having excess BMI. Compared to Veterans who died without progression or were censored, Veterans who progressed to MM were younger at MGUS diagnosis (median: 67.6 (progression) versus 75.0 (died) versus 69.9 (censored) years, p<0.0001), had shorter follow-up (median: 2.3 versus 3.7 versus 4.3 years, p<0.0001), were more likely to be Black (39.1% versus 29.3% versus 37.0%, p<0.0001), have abnormal M-spike level at study entry (24.4% versus 8.4% versus 4.2%, p<0.0001), and have IgA subtype (17.7% versus 11.9% versus 11.5%, p<0.0001).

**Table 1.** Characteristics by progression status in Veterans diagnosed with MGUS in the VHA from 10/1/1999 to 12/31/2023.

|  | Total | Progression | Death | Censored | p-value* |
| --- | --- | --- | --- | --- | --- |
| **No.** | 35,073 | 2,895 | 15,541 | 16,637 |  |
| **%** | 100.0% | 8.3% | 44.3% | 47.4% |  |
| **Gender, No. (%)** |  |  |  |  | <0.0001 |
| Female | 1,403 (4.0) | 90 (3.1) | 295 (1.9) | 1,031 (6.2) |  |
| Male | 33,670 (96.0) | 2,805 (96.9) | 15,246 (98.1) | 15,606 (93.8) |  |
| **Race, No. (%)** |  |  |  |  | <0.0001 |
| Black | 11,855 (33.8) | 1,132 (39.1) | 4,554 (29.3) | 6,156 (37.0) |  |
| White | 23,218 (66.2) | 1,763 (60.9) | 11,003 (70.8) | 10,481 (63.0) |  |
| **Excess BMI, No. (%)** |  |  |  |  | <0.0001 |
| Non-excess BMI | 9,119 (26.0) | 620 (21.4) | 4,973 (32.0) | 3,510 (21.1) |  |
| Excess BMI | 25,638 (73.1) | 2,264 (78.2) | 10,412 (67.0) | 12,977 (78.0) |  |
| Missing | 316 (0.9) | 14 (0.5) | 155 (1.0) | 150 (0.9) |  |
| **BMI, No. (%)** |  |  |  |  | <0.0001 |
| Underweight | 666 (1.9) | 23 (0.8) | 435 (2.8) | 183 (1.1) |  |
| Normal weight | 8,453 (24.1) | 596 (20.6) | 4,538 (29.2) | 3,327 (20.0) |  |
| Overweight | 12,451 (35.5) | 1,062 (36.7) | 5,470 (35.2) | 5,939 (35.7) |  |
| Obese | 13,187 (37.6) | 1,201 (41.5) | 4,942 (31.8) | 7,037 (42.3) |  |
| Missing | 316 (0.9) | 14 (0.5) | 155 (1.0) | 150 (0.9) |  |
| **M-spike, No. (%)** |  |  |  |  | <0.0001 |
| 0 | 1,964 (5.6) | 128 (4.4) | 755 (4.9) | 1,080 (6.5) |  |
| Not quantifiable | 4,395 (12.5) | 353 (12.2) | 2,129 (13.7) | 1,913 (11.5) |  |
| ≤1.5 g/dL | 22,882 (65.2) | 1,464 (50.6) | 9,782 (62.9) | 11,636 (69.9) |  |
| >1.5 g/dL | 2,718 (7.8) | 706 (24.4) | 1,310 (8.4) | 704 (4.2) |  |
| Missing | 3,114 (8.9) | 244 (8.4) | 1,565 (10.1) | 1,304 (7.8) |  |
| **MGUS type, No. (%)** |  |  |  |  | <0.0001 |
| IgA | 4,279 (12.2) | 512 (17.7) | 1,849 (11.9) | 1,913 (11.5) |  |
| IgG | 24,937 (71.1) | 2,058 (71.1) | 10,941 (70.4) | 11,945 (71.8) |  |
| Light-chain | 5,857 (16.7) | 324 (11.2) | 2,735 (17.6) | 2,778 (16.7) |  |
| **Chemical exposure, No. (%)** |  |  |  |  | <0.0001 |
| No | 31,531 (89.9) | 2,553 (88.2) | 14,453 (93.0) | 14,507 (87.2) |  |
| Yes | 3,542 (10.1) | 342 (11.8) | 1,088 (7.0) | 2,130 (12.8) |  |
| **CCI↑, No. (%)** |  |  |  |  | <0.0001 |
| 0 | 5,454 (15.6) | 391 (13.5) | 1,582 (10.2) | 3,482 (20.9) |  |
| >0 | 29,619 (84.5) | 2,504 (86.5) | 13,959 (89.8) | 13,155 (79.1) |  |
| **Follow-up (years)** |  |  |  |  | <0.0001 |
| Median | 3.8 | 2.3 | 3.7 | 4.3 |  |
| IQR | 1.5-7.3 | 1.1-4.9 | 1.5-7.0 | 1.7-8.0 |  |
| **Age** |  |  |  |  | <0.0001 |
| Median | 71.8 | 67.6 | 75.0 | 69.9 |  |
| IQR | 64.4-78.6 | 60.9-74.2 | 67.4-82.0 | 62.6-75.7 |  |
| **Calendar year of MGUS diagnosis** |  |  |  |  | <0.0001 |
| Median | 2016 | 2012 | 2012 | 2019 |  |
| IQR | 2010-2020 | 2007-2017 | 2007-2017 | 2015-2022 |  |

MGUS, monoclonal gammopathy of undetermined significance; MM, multiple myeloma; aHR, adjusted hazard ratio; aPAF, adjusted population attributable fraction; CI, confidence interval; Ig, immunoglobulin; CCI, Charlson comorbidity index; IQR, interquartile range.
\* Chi-square tests were conducted to compare percentages for categorical variables and Kruskal–Wallis tests were used to compare medians for continuous variables between the three groups.
↑ CCI was calculated based on data collected within 1 year before MGUS diagnosis.

Patients across racial groups were statistically significantly different in all variables (eTable 1).

### Competing risk analyses

The cumulative incidence of progression varied significantly across categories of each modifiable risk factor (Figure 1A-C). Notably, the cumulative incidence of progression was significantly higher for patients who had excess BMI (p<0.0001), chemical exposure (p<0.0001), comorbidities >0 (p=0.0009).

Multivariable analyses demonstrated significant associations with the progression of MGUS to MM for all included modifiable factors (Table 2): excess BMI (aHR=1.24, 95% CI=1.13-1.36), chemical exposure (aHR=1.29, 95% CI=1.15-1.45), and comorbidities (aHR=1.34, 95% CI=1.20-1.49).

**Table 2.** Overall and race-specific multivariable-adjusted hazard ratios for the progression of MGUS to MM.

| Risk factors | All |  | White |  | Black |  |
| --- | --- | --- | --- | --- | --- | --- |
|  | aHR (95% CI)* | p-value | aHR (95% CI)* | p-value | aHR (95% CI)* | p-value |
| Age | 0.970 (0.967-0.974) | <.0001 | 0.966 (0.962-0.970) | <.0001 | 0.977 (0.972-0.982) | <.0001 |
| Gender |  |  |  |  |  |  |
| Female | 1.00 (referent) |  | 1.00 (referent) |  | 1.00 (referent) |  |
| Male | 1.49 (1.20-1.85) | 0.0003 | 1.64 (1.18-2.28) | 0.0031 | 1.35 (1.02-1.80) | 0.0386 |
| M-spike<sup>†</sup> |  |  |  |  |  |  |
| ≤1.5 g/dL | 1.00 (referent) |  | - |  | - |  |
| >1.5 g/dL | 1.12 (1.03-1.21) | 0.0052 | - | - | - | - |
| Race |  |  |  |  |  |  |
| White | 1.00 (referent) |  | 1.00 (referent) |  | 1.00 (referent) |  |
| Black | 4.38 (3.99-4.81) | <.0001 | 4.61 (4.08-5.20) | <.0001 | 4.12 (3.56-4.77) | <.0001 |
| Ig subtype<sup>†</sup> |  |  |  |  |  |  |
| IgG | 1.00 (referent) |  | 1.00 (referent) |  | 1.00 (referent) |  |
| IgA | 1.58 (1.43-1.74) | <.0001 | 1.72 (1.52-1.95) | <.0001 | 1.39 (1.18-1.63) | <.0001 |
| Light-chain | 0.67 (0.58-0.78) | <.0001 | 0.63 (0.53-0.76) | <.0001 | 0.76 (0.60-0.97) | 0.0263 |
| BMI<sup>†</sup> |  |  |  |  |  |  |
| Non-excess BMI | 1.00 (referent) |  | 1.00 (referent) |  | 1.00 (referent) |  |
| Excess BMI | 1.24 (1.13-1.36) | <.0001 | 1.20 (1.07-1.35) | 0.0023 | 1.30 (1.13-1.50) | 0.0003 |
| Chemical exposure |  |  |  |  |  |  |
| No | 1.00 (referent) |  | 1.00 (referent) |  | 1.00 (referent) |  |
| Yes | 1.29 (1.15-1.45) | <.0001 | 1.38 (1.20-1.59) | <.0001 | 1.09 (0.89-1.34) | 0.3952 |
| CCI |  |  |  |  |  |  |
| 0 | 1.00 (referent) |  | 1.00 (referent) |  | 1.00 (referent) |  |
| >0 | 1.34 (1.20-1.49) | <.0001 | 1.37 (1.20-1.57) | <.0001 | 1.28 (1.07-1.52) | 0.0065 |
| Year of MGUS diagnosis | 0.98 (0.97-0.98) | <.0001 | 0.97 (0.96-0.98) | <.0001 | 0.98 (0.97-0.99) | 0.0016 |

MGUS, monoclonal gammopathy of undetermined significance; MM, multiple myeloma; aHR, adjusted hazard ratio; CI, confidence interval; Ig, immunoglobulin; CCI, Charlson comorbidity index.
\*HRs were additionally adjusted for M-spike level at MGUS (0, not quantifiable, ≤1.5 g/dL, >1.5 g/dL, missing), and year of MGUS diagnosis.
<sup>†</sup> Missing M-spike, and missing BMI categories are not presented.

### aPAF analyses

Excess BMI was responsible for 27.1% (95% CI=22.1-31.9%) of MM cases in the VHA (Figure 2A). Absence of AO exposure may have reduced the MM burden by 3.9% (95% CI=2.5-5.2%). If all patients had no comorbidities, the MM burden may have been 8.0% higher, although not statistically significant (95% CI=-16.7-0.1%). If all patients had ≥1comorbidity (reverse comorbidity from CCI=0 to >0), the MM burden may have been 0.9% lower (95% CI=-0.8-2.6%). See eTable 2 for all risk factors. Age of MGUS diagnosis decreased by one-year, male gender, black race, and IgA subtype accounted for 5.1% (95% CI=4.9-5.4%), 15.1% (95% CI=-2.1-29.4%), 5.2% (95% CI=2.5-7.8%), and 5.2% (95% CI=3.9-6.6%) of MM cases, respectively.

**Figure 2.**
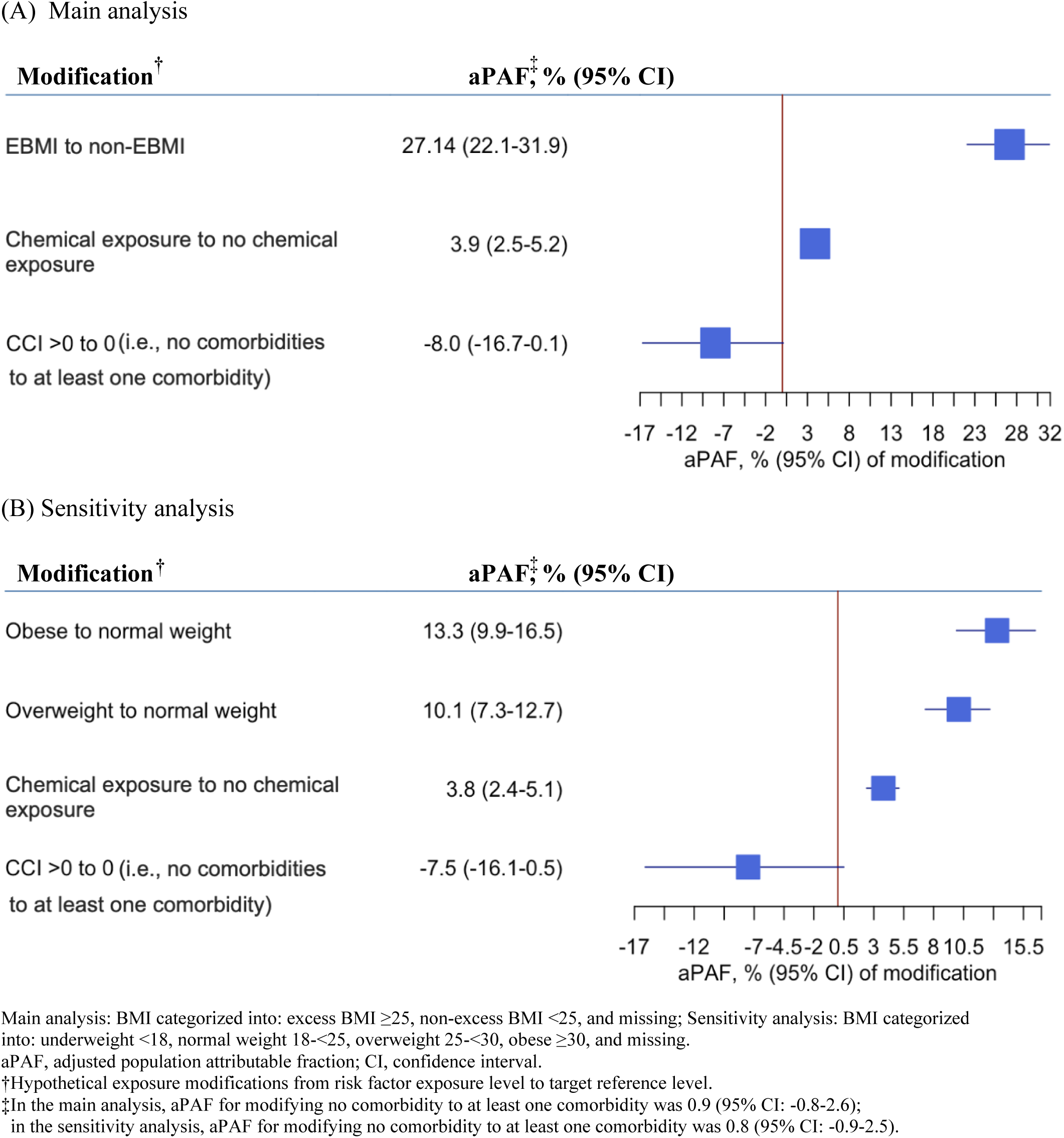
Multivariable-adjusted population attributable fractions for selected modifiable risk factors of the progression of MGUS to MM.

### Race-specific analyses

Excess BMI was associated with an elevated progression risk in both White (aHR=1.20, 95% CI=1.07-1.35) and Black patients (aHR=1.30, 95% CI=1.13-1.50) (Table 2). Having comorbidities was associated with a higher risk of MGUS progression in both groups (aHR_White_=1.37, 95% CI=1.20-1.57; aHR_Black_=1.28, 95% CI=1.07-1.52). Chemical exposure was significantly associated with a higher progression risk only for White patients (aHR_White_=1.38, 95% CI=1.20-1.59; aHR_Black_=1.09, 95% CI=0.89-1.34).

Excess BMI emerged as the top modifiable risk factor across both racial groups (Figure 3A and eTable 2, aPAF_White_=27.2%, 95% CI=20.3-33.4%; aPAF_Black_=27.1%, 95% CI=19.5-34.0%; p-difference=0.9806,). The contribution of chemical exposure was greater for White patients (aPAF_White_=5.6%, 95% CI=3.6-7.6%; aPAF_Black_=1.7%, 95% CI=0.1-3.3%; p-difference=0.0027). Changing CCI from >0 to 0 increased in MM cases 9.7% among White patients (95% CI=-21.0-0.6%) and 5.8% among Black patients (95% CI=-19.6-6.5%) (p-difference=0.4865). See eTable 2 for race-specific aPAFs for all risk factors.

**Figure 3.**
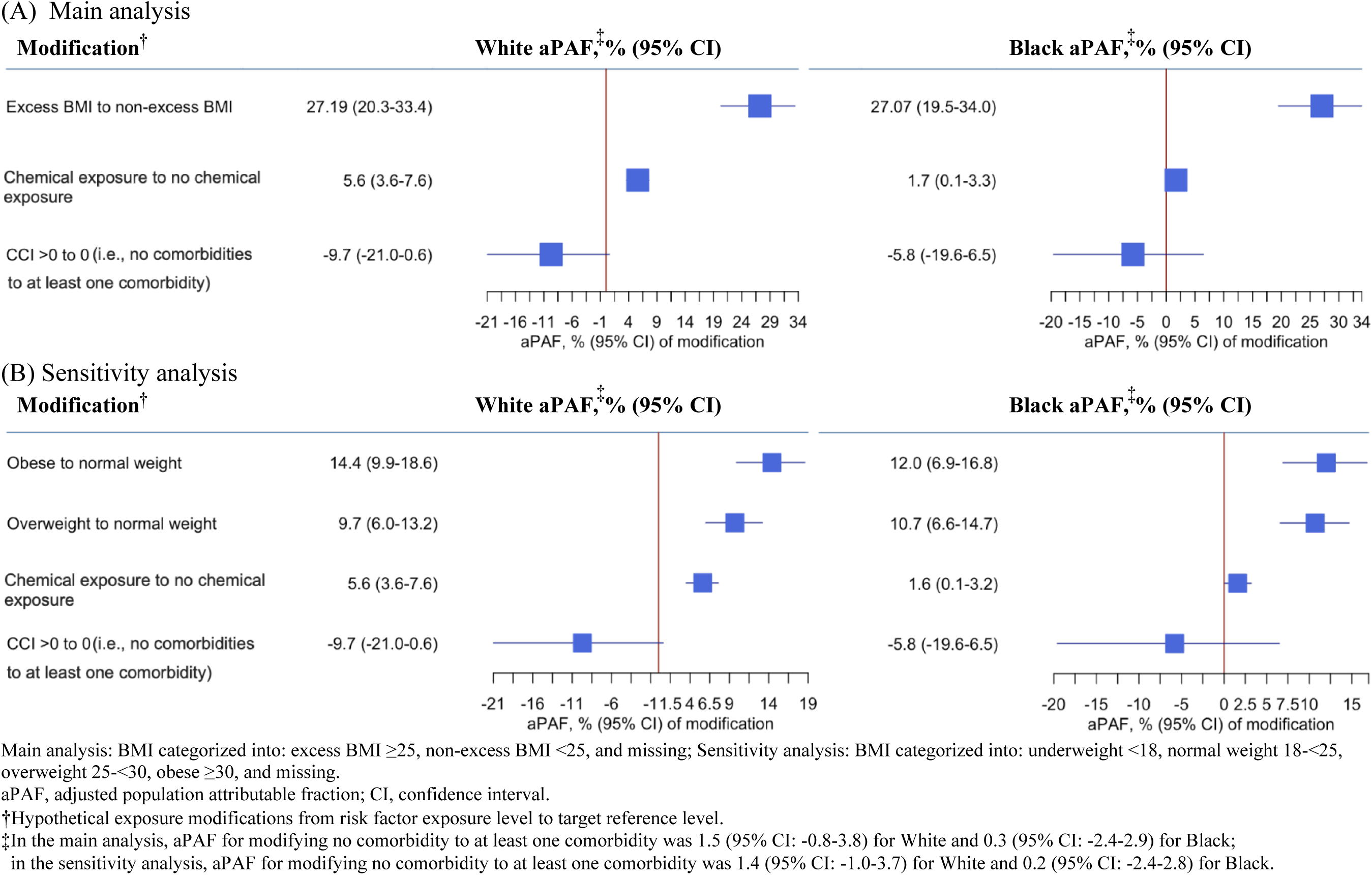
Race-specific multivariable-adjusted population attributable fractions for selected modifiable risk factors of the progression of MGUS to MM.

### Sensitivity Analyses

Obesity and overweight were the top two modifiable risk factors (Figure 2B, aPAF_obesity_=13.3%, 95% CI=9.9-16.5%; aPAF_overweight_=10.1%, 95% CI=7.3-12.7%). The burden of MM cases attributable to obesity was higher for White patients (aPAF_White_=14.4%, 95% CI=9.9-18.6%), compared to Black patients (aPAF_Black_=12.0%, 95% CI=6.9-16.8%) (Figure 3B). The proportion of MM cases attributable to overweight was lower for White than Black patients (aPAF_White_=9.7%, 95% CI=6.0-13.2%; aPAF_Black_=10.7%, 95% CI=6.6-14.7%).

## DISCUSSION

To our knowledge, this is the first study to quantify the burden of MGUS progression attributable to each identified risk factor. In this national cohort of Veterans, excess BMI was the most significant contributor to the progression of MGUS to MM, accounting for 27.1% of the progression cases, consistently observed across both racial groups: White 27.2%; Black 27.1%.

Our study highlights the crucial role of obesity in MGUS progression, supported by prior research indicating that obesity promotes the progression through several biological mechanisms.^7,19,55–57^ Obesity is linked to elevated levels of inflammatory cytokines, including TNF-α, CRP, and IL-6, which support MM cell survival and proliferation.^55,58,59^ Weight loss has been shown to significantly reduce these cytokines.^60–62^ Additionally, obesity disrupts adipokine balance, increasing leptin and decreasing adiponectin, further influencing MGUS progression.^63,64^ Given its strong contribution to risk and modifiability, weight management may mitigate the progression of MGUS to MM.

Comorbidities, including type-2 diabetes mellitus and other obesity-related diseases (e.g., hypertension, cardiovascular disease) were linked to increased progression risk based on CIF and multivariable analysis. However, the aPAF for modifying comorbidity was not statistically significant. Both progression and death were considered for estimating the aPAFs for progression. A higher CCI is associated with both outcomes, and reducing CCI from >0 to 0 may reduce more deaths than progression events, potentially increasing observed progression cases. This relationship may be further complicated by shared metabolic and inflammatory mechanisms with obesity.

Our study also quantified the contribution of chemical herbicide exposure, which accounted for 3.9% of MM cases. Previous studies have identified chemical exposure, possibly due TCDD contamination, as a risk factor for MGUS and its progression.^39–43^ TCDD is common in various occupational pesticides. The VHA data provide a unique opportunity to study the impact of chemical exposure via the exposure of AO, offering insights into the long-term effects of environmental and occupational chemical exposures and informing policies for exposure prevention and management. Similar studies have found that occupational pesticide exposure is associated with higher MGUS and MM risk.^13–17^ Therefore, our findings underscore the importance of considering environmental exposures in MM prevention, particularly for MGUS patients at high risk due to occupational or environmental TCDD exposure.

We found that younger age at MGUS diagnosis was associated with a higher risk of progression to MM, aligning with previous studies.^42,65^ Similar to comorbidities, this negative association may stem from older age being strongly associated with higher all-cause mortality risk. Older individuals with shorter lifespans post-MGUS diagnosis may not live long enough for MGUS to progress to MM,^8^ resulting in a negative association with progression risk. Our data also showed that patients diagnosed with MGUS at an older age (≥ median age of 71.8 years) had a shorter average follow-up duration (3.9 years), compared to younger patients (<71.8 years) who had 6.1 years. This difference may be attributed to higher mortality rates in the older population (54.4%) versus the younger cohort (34.2%). Therefore, it is crucial to investigate whether shorter follow-up times or comorbidities in older patients influence the progression of MGUS to MM.

While previous research has indicated increased risk of MGUS and MM among Black compared to White populations,^26,28,31^ our race-specific analyses showed larger aPAFs for selected modifiable risk factors in White compared to Black individuals. These disparities may be attributed to socio-contextual factors, potentially linked to underlying health inequities.^66,67^ Excess BMI emerged as a prominent modifiable risk factor for MGUS progression in both racial groups (aPAF_White_: 27.2%; aPAF_Black_: 27.1%). Although this difference was not statistically significant, addressing excess BMI as a modifiable risk factor could substantially reduce MM burden in both racial groups.

We found that the contribution of chemical exposure was more pronounced in White compared to Black individuals, and no statistically significant racial difference existed in the contribution of modifying comorbidities from 0 to ≥1. In our cohort, the prevalence of chemical exposure was slightly higher in White than in Black individuals. Multivariable competing-event analyses showed that the impact of chemical exposure was greater in White compared to Black individuals. These findings warrant further studies on racial differences in the impact of chemical exposure on MGUS progression and development of equitable, population-specific intervention strategies.

In addition to the analyzed modifiable risk factors, factors like family history/genetics, might contribute to the observed racial differences in MM progression,^67^ necessitating further research to elucidate the underlying mechanisms of these findings and to explore potential contributing factors. Ultimately, these results suggest the need to develop tailored interventions for specific subpopulations.^6,31,67,68^

Our study has several strengths. First, we used a large national Veterans cohort with NLP-confirmed MGUS/MM diagnoses, improving diagnosis accuracy from 20% to 93% for MGUS and from 58% to 99% for MM compared to administrative codes alone.^44–46^ Second, we leveraged the well-documented chemical exposure data of the Veterans population to investigate the association of chemical exposure on MGUS progression at the population level. The detailed VHA data allowed a more precise assessment of this usually hard-to-capture risk factor. Third, race-specific aPAFs analysis explored potential racial differences in the contribution of modifiable risk factors to MGUS progression. Identifying higher burden racial group helps in planning targeted prevention strategies to reduce health disparities. As racial disparities in MM are long-established, examining the contribution of modifiable risk factors separately for White and Black populations provides insight into any differential contribution of these factors on MM development, potentially identifying interventions to reduce MM health disparities. Fourth, the application of a comprehensive aPAF method that accounts for the competing risk of death addresses a crucial limitation in studies examining disease progression; specifically, MGUS/MM predominantly affect older adults who face a higher risk of dying from causes unrelated to the disease of interest before progressing to the disease outcome. This enhances the validity of our findings by minimizing the possibility of overestimating aPAF.

Our study has limitations. First, risk factors like family history,^11,68,69^ smoking status,^11,70^ and dietary intake^71^ are not recorded as structured data in the VHA. Further data abstraction or development of NLP algorithms is needed to retrieve these data. Second, the male-dominant Veterans population limits the generalizability of our findings to female populations. However, the insights gained from this population can inform future investigation in more diverse cohorts. Third, the retrospective design precludes establishing causality between risk factors and MGUS progression. Fourth, despite using a validated NLP model to confirm MGUS and MM diagnoses, misclassification cannot be completely ruled out. However, we do not anticipate that the misclassification is high as our study shows that the NLP model achieved high accuracy (MGUS: 93%; MM: 99%). Last, the interpretation of aPAFs should focus on the contribution of these risk factors to the progression of MGUS to MM rather than the practical intervention impact, as modifying these risk factors in practice may be complex. Despite these limitations, our study offers insights into the relative contributions of key risk factors in MGUS progression to MM among Veterans.

## CONCLUSION

Our study quantifies the relative contribution of modifiable risk factors to the progression of MGUS to MM among Veterans. Among the variables studied, our results indicate that excess BMI is the top contributor. As one of the few modifiable risk factors identified, these results underscore the potential impact of targeted interventions like lifestyle modifications and weight management on reducing MGUS progression. Further research is needed to validate these findings across different populations and to identify additional risk factors to inform MM prevention.

## Data Sharing Statement

The data obtained from the VHA for the analyses in this study contain patient health information, including Social Security numbers and addresses. The institutional review board (IRB)–approved study protocol allows us to share analysis results only in an aggregate format.

We also have the following policies in place that do not allow us to share the data: In our **original IRB application** under “Data Management and Access Plan”:

Will final data sets underlying the publications be shared outside the US Department of Veterans Affairs (VA)

No. The final data sets contain patient health information and will not be shared outside of the VA.

In our **data use agreement** with VA Information Resource Center for use of VA and Centers for Medicare & Medicaid Services (CMS) data:

**Data security**: VA/CMS data will be protected in accordance with the project’s VA/CMS Data Security Compliance Form and may be stored and used only on VA Office of Information and Technology–managed network servers located at: VA Facility: VA Austin Information Technology Center, Austin, Texas VA Informatics and Computing Infrastructure workspace: ORD_Chang_202011003D

In our data access permissions:

Access to these data will be restricted to only VA-approved project employees whose roles and responsibilities necessitate access to these data (hereinafter referred to as authorized data users) and who have signed and submitted a VA/CMS Rules of Behavior Agreement.

## Data Availability

The data obtained from the VHA for the analyses in this study contain patient health information, including Social Security numbers and addresses. The institutional review board (IRB) approved study protocol allows us to share analysis results only in an aggregate format. Access to these data will be restricted to only VA-approved project employees whose roles and responsibilities necessitate access to these data (hereinafter referred to as authorized data users) and who have signed and submitted a VA/CMS Rules of Behavior Agreement.

