## Supplemental Figure 1, Table 1, Table 2 for "Quantify the Contribution of Modifiable Risk Factors for Progression of MGUS to Multiple Myeloma"

Mei Wang, MS[1,2,3†]; Byron Sigel, MsC,[4†]; Lawrence Liu, MD[5]; John H. Huber, PhD[3]; Mengmeng Ji, PhD, MBBS[1,3]; Martin W. Schoen, MD, MPH[1,6]; Kristen M. Sanfilippo, MD, MPHS[1,4]; Theodore S. Thomas, MD, MPHS[1,4]; Graham A. Colditz, MD, DrPH[3], Shi-Yi Wang, MD, PhD[7]; Su-Hsin Chang, PhD, SM[1,3*]
1. Research Service, St. Louis Veterans Affairs Medical Center; St. Louis, MO
2. Division of Biology and Biomedical Sciences, Washington University School of Medicine; St. Louis, MO
3. Division of Public Health Sciences, Department of Surgery, Washington University School of Medicine; St. Louis, MO
4. Division of Hematology and Medical Oncology, Department of Internal Medicine, Saint Louis University School of Medicine, St. Louis, MO
5. City of Hope Comprehensive Cancer Center; Duarte, CA
6. Department of Medicine, Washington University School of Medicine; St. Louis, MO
7. Yale School of Public Health, Yale University; New Haven, CT
[†]Co-first authors.

*Corresponding Author: Su-Hsin Chang, PhD SM

Address: 660 S. Euclid Ave. Campus Box 8100, St. Louis, MO 63110

Funding: This work was supported by the Foundation for Barnes-Jewish Hospital; the Siteman Cancer Center; the National Institutes of Health Grants R01 CA253475 and U01 CA265735.

eFigure 1. Flowchart of patient selection
eTable 1. Characteristics by race in Veterans diagnosed with MGUS in the VHA from 10/1/1999 to 12/31/2023
eTable 2. Multivariable-adjusted population attributable fractions for selected risk factors of the progression of MGUS to MM.

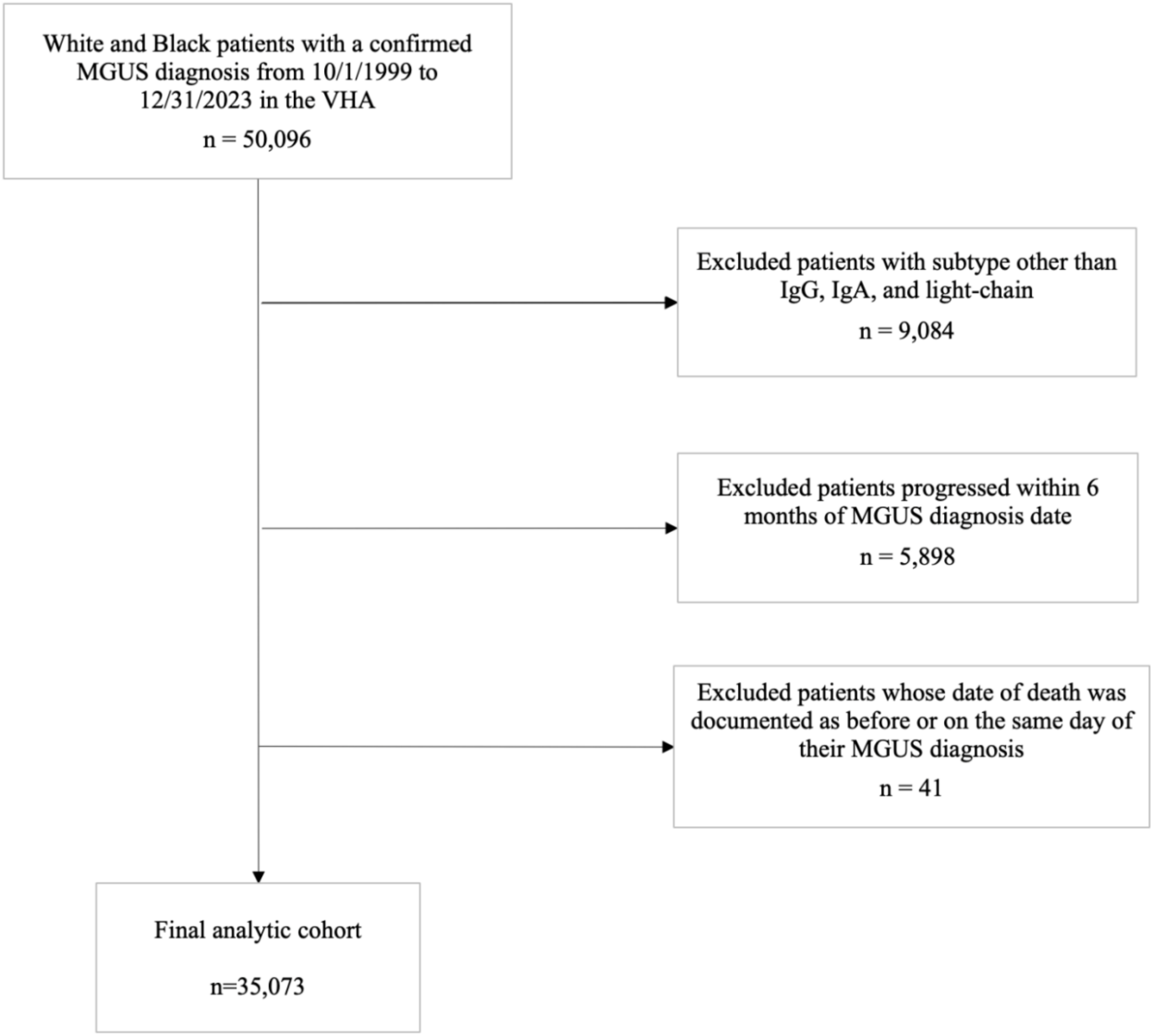


eFigure 1. Flowchart of patient selection

eTable 1. Characteristics by race in Veterans diagnosed with MGUS in the VHA from 10/1/1999 to 12/31/2023.

| | White | Black | P-value* |
|---|---|---|---|
| **N** | 23,234 | 11,839 | |
| **%** | 66.2% | 33.8% | |
| **Outcome (%)** | | | <0.0001 |
| Confirmed progression | 7.6 | 9.6 | |
| Death without progression | 47.3 | 38.4 | |
| Censoring | 45.1 | 52.0 | |
| **Gender (%)** | | | <0.0001 |
| Female | 3.1 | 5.9 | |
| Male | 96.9 | 94.1 | |
| **Excess BMI (%)** | | | <0.0001 |
| Non-excess BMI | 24.8 | 28.3 | |
| Excess BMI | 74.2 | 71.0 | |
| Missing | 1.0 | 0.7 | |
| **BMI (%)** | | | <0.0001 |
| Underweight | 1.5 | 2.6 | |
| Normal weight | 23.3 | 25.6 | |
| Overweight | 36.7 | 33.4 | |
| Obese | 37.5 | 37.7 | |
| Missing | 1.0 | 0.7 | |
| **M-spike (%)** | | | <0.0001 |
| 0 | 5.45 | 5.88 | |
| Not quantifiable | 12.39 | 12.81 | |
| ≤1.5 g/dL | 64.17 | 67.35 | |
| >1.5 g/dL | 7.39 | 8.46 | |
| Missing | 10.6 | 5.5 | |
| **MGUS type (%)** | | | <0.0001 |
| IgA | 11.6 | 13.4 | |
| IgG | 69.8 | 73.8 | |
| Light-chain | 18.7 | 12.8 | |
| **Chemical exposure (%)** | | | <0.0001 |
| No | 89.0 | 91.6 | |
| Yes | 11.0 | 8.4 | |
| **CCI↑ (%)** | | | <0.0001 |
| 0 | 94.4 | 95.3 | |
| >0 | 5.6 | 4.7 | |
| **Age (years)** | | | <0.0001 |
| Median | 73.3 | 68.4 | |
| IQR | 66.3-79.8 | 61.0-75.6 | |

| | | | |
|---|---|---|---|
| **Follow-up (years)** | | | <0.0001 |
| Median | 3.7 | 4.0 | |
| IQR | 1.5-7.2 | 1.6-7.6 | |
| **Calendar year of MGUS diagnosis** | | | 0.0384 |
| Median | 2016 | 2016 | |
| IQR | 2010-2020 | 2011-2020 | |

MGUS, monoclonal gammopathy of undetermined significance; MM, multiple myeloma; aHR, adjusted hazard ratio; aPAF, adjusted population attributable fraction; CI, confidence interval; Ig, immunoglobulin; CCI, Charlson comorbidity index; IQR, interquartile range.

* Chi-square tests were conducted to compare percentages for categorical variables and Kruskal–Wallis tests were used to compare medians for continuous variables between the three groups.

↑ CCI was calculated based on data collected within 1 year before MGUS diagnosis.

eTable 2. Multivariable-adjusted population attributable fractions for selected risk factors of the progression of MGUS to MM.

| Risk factors | Modifications† | aPAF, % (95% CI)* | | | |
|---|---|---|---|---|---|
| | | Overall | White | Black | p-difference** |
| Age | | | | | |
| | Increase by 1 year | 5.1 (4.9-5.4) | 5.8 (5.5-6.1) | 4.3 (3.9-4.7) | <0.0001 |
| Gender | | | | | |
| Female | Male to Female | 15.1 (-2.1-29.4) | 24.8 (-0.9-43.9) | 4.2 (-20.1-23.6) | 0.1878 |
| Male | | | | | |
| Race | | | | | |
| White | Black to White | 5.2 (2.5-7.8) | - | - | - |
| Black | | | | | |
| Ig subtype[↑] | | | | | |
| IgG | IgA to IgG | 5.2 (3.9-6.6) | 6.0 (4.3-7.7) | 4.2 (2.1-6.3) | 0.2077 |
| | Light-chain to IgG | -7.0 (-8.7--5.3) | -8.2 (-10.9--5.9) | -5.5 (-7.4--3.6) | 0.0473 |
| IgA | | | | | |
| Light-chain | | | | | |
| Excess BMI status[↑] | | | | | |
| Non-excess BMI | Excess BMI to non-excess BMI | 27.14 (22.1-31.9) | 27.19 (20.3-33.4) | 27.07 (19.5-34.0) | 0.9806 |
| Excess BMI | | | | | |
| CCI | | | | | |
| | >0 to 0 (i.e., at least one comorbidity to no comorbidities) | 1.0 (-0.8-2.7) | 1.5 (-0.8-3.8) | 0.3 (-2.4-2.9) | 0.4865 |
| Chemical exposure | | | | | |
| No | Chemical exposure to No chemical exposure | 3.9 (2.5-5.2) | 5.6 (3.6-7.6) | 1.7 (0.1-3.3) | 0.0027 |
| Yes | | | | | |

MGUS, monoclonal gammopathy of undetermined significance; MM, multiple myeloma; aHR, adjusted hazard ratio; aPAF, adjusted population attributable fraction; CI, confidence interval; Ig, immunoglobulin; CCI, Charlson comorbidity index.

†Hypothetical exposure modifications from risk factor exposure level to target reference level.

*HRs and aPAFs were additionally adjusted for M-spike level at MGUS (0, not quantifiable, ≤1.5 g/dL, >1.5 g/dL, missing), and year of MGUS diagnosis.

**p-differences were based on two-sample z-tests in comparing differences in aPAFs between White and Black.

↑Missing BMI category is not presented.
